# Breast Cancer Survival Prediction using Machine Learning and Gene Expression Profiles

**DOI:** 10.1101/2022.01.22.22269470

**Authors:** Dheiver Santos

## Abstract

Breast cancer is one of the most common cancers with a high mortality rate among women. It is also the most frequent cause of cancer death in this population, with an estimated 684,996 deaths for that year (15.5% of cancer deaths in women) (IARC, 2020). In Brazil, breast cancer is also the most common type of cancer in women from all regions, after non-melanoma skin cancer. Therefore, an accurate and reliable system is needed for the early diagnosis of this cancer. Machine learning explores the study and construction of algorithms that can learn from their mistakes and make predictions about data. The results here are excellent with an accuracy and precision level of around 79%.

## 1. Introduction

Cancer, also known as neoplasm or malignant tumor, is considered one of the major problems in the world of health. About 70% of deaths caused by cancer in general occur in low- and middle-income countries, but approximately 30% of the total number of deaths due to cancer are preventable [1] However, studies by the International Agency for Research on Cancer report that the number of deaths caused by cancer will continue to increase, with an estimated 12 million deaths by mid-2030 [2]. When specifically referring to breast cancer, it corresponds to 22% of the types of cancer detected annually and that it has a higher incidence in women than in men, being considered the most frequent type of cancer in the world and the most common type of cancer. first that leads to the death of female individuals [3,4,5] In Brazil, the National Cancer Institute (INCA) reports that in 2014 there were 57,124,000 new cases of breast cancer, an average of 57 cases per 100,000 women. As for the number of deaths in the same year, it was 13,345 occurrences. It is worth noting that despite the high mortality rate currently found, breast cancer is considered a disease with a good prognosis [6,7]

In recent decades, a heightened interest in the use of pattern recognition techniques, processing, analysis of genetics and artificial intelligence has emerged around the world. The set of these techniques has been constantly used for the development of detection support systems (CADe) and diagnostic support systems (CADx) [6,7]. The objective of this work is to develop a system to support the diagnosis of breast cancer by analyzing genetics from classifiers based on Machine Learning Boost and statistical attributes, with the application of attribute selection techniques, and focus on an algorithm. supervised.

## 2. Database

In a predictive model is developed for the evaluation of survival in women that have been diagnosed with breast cancer, while they addressed the importance of robustness under the model’s parameter variation.

The Molecular Taxonomy of Breast Cancer International Consortium (METABRIC) database is a Canada-UK Project which contains targeted sequencing data of 1,980 primary breast cancer samples. Clinical and genomic data was downloaded from cBioPortal.

The dataset was collected by Professor Carlos Caldas from Cambridge Research Institute and Professor Sam Aparicio from the British Columbia Cancer Centre in Canada and published on Nature Communications (Pereira et al., 2016) [1]. It was also featured in multiple papers including Nature and others:

> *Associations between genomic stratification of breast cancer and centrally reviewed tumour pathology in the METABRIC cohort*
>
> *Predicting Outcomes of Hormone and Chemotherapy in the Molecular Taxonomy of Breast Cancer International Consortium (METABRIC) Study by Biochemically-inspired Machine Learning*

### Genetic attributes in the dataset

The genetics part of the dataset contains m-RNA levels z-score for 331 genes, and mutation for 175 genes.

## 3. Machine Learning

The methodology of this work is an adaptation of the CRISP-DM methodology, and was divided into three stages: data extraction; selection and pre-processing of data; execution and optimization of hyperparameters of the XGBoost algorithm to obtain the ranking results.

This work was developed using the Python language, together with the Scikit-Learn module and the XGBoost libraries, which implement the tree boosting algorithm, and Hyperopt responsible by optimizing the hyperparameters.

To carry out the experiments, the k-fold cross-validation method of 10 segments. For this purpose, the data was also partitioned into 10 segments or folds. Therefore, for each execution, the data of each segment was distributed as follows:

In these folds, training and testing are done in 10 iterations, so that in eachiteration, model training is done with 9 segments and testing is done on the remaining fold. The evaluation metrics are obtained in each iteration, and at the end of the iterations, mean and standard deviation are calculated, in order to evaluate the model.

One The schematic of this process is shown in Figure 16, which shows the division of data into training and testing, model validation with validation data and metrics extraction.

Step by Step

> *Six Easy Steps to build XGBoost model in python:*
>
> *Step 1: Install libraries, xgboost, margrittr, Matrix*
>
> *Step 2: Create a Matrix for train and test datasets-with the use of xgb*.*DMatrix() function*
>
> *Step 3: Set parameters, for params and watchlist*
>
> *Step 4: Build model using xgb*.*train() function*
>
> *Step 5: Use xgb*.*importance() function for feature analysis*
>
> *Step 6: Make prediction with the use of predict() function*

Next, will be presented and discussed the results obtained with the application of the proposed methodology.

## 4. Results

For the distribution of all numerical data (Figure 1), some of them are normally distributed (like tumor_stage, and age_at_diagnosis), but most of the features are right skewed with a lot of outliers (lymph_nodes_examined_positive, mutation_count, and tumor_size). We decided to keep the outliers, as they are very important in healthcare data.

**Figure 1.**
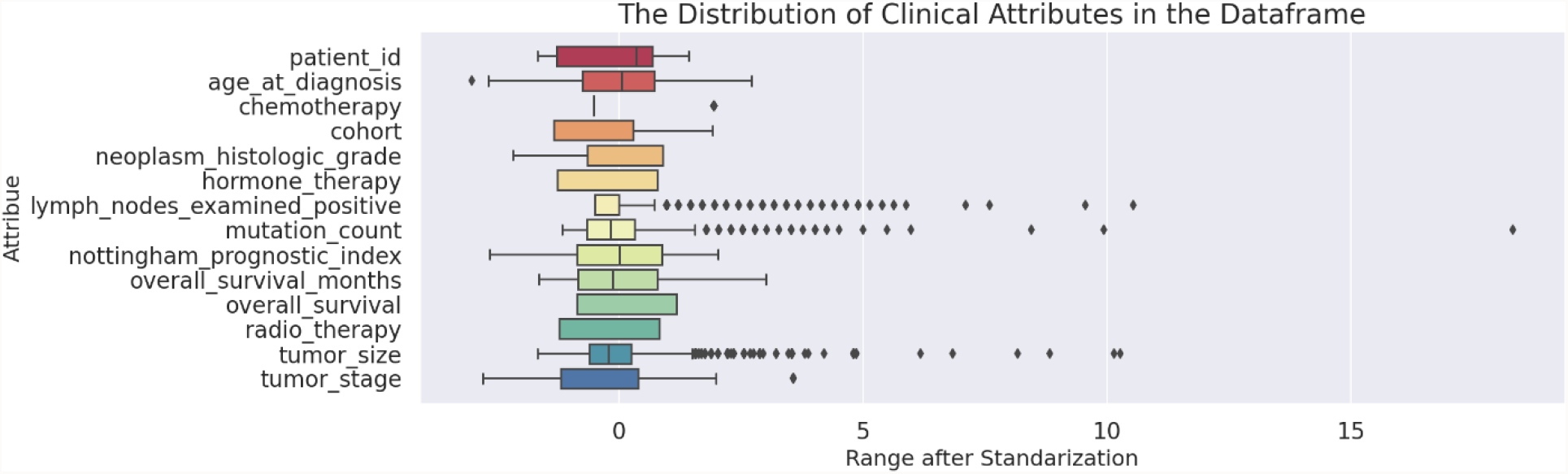
BoxPlot Dataset

it is very important to assess whether the dataset contains significant values of outliers, large bars mean significant variations within the variable, small bars mean small variations and it is also important to understand where this average is in relation to a given variable, such as size of tumor.

The Figure 2 presents a probability distribution, for example, in relation to the size of the tumor, it is possible to see that the average, when we talk about the size of the tumor, is between 0 and 50 and that in this range the distributions overlap, in this range you can see that the distributions are almost identical in relation to people who survived and who are alive, in relation to people who died, this analysis of probability distribution in variables is very important to give an overview with respect to the database of living and dead people and its implications

**Figure 2.**
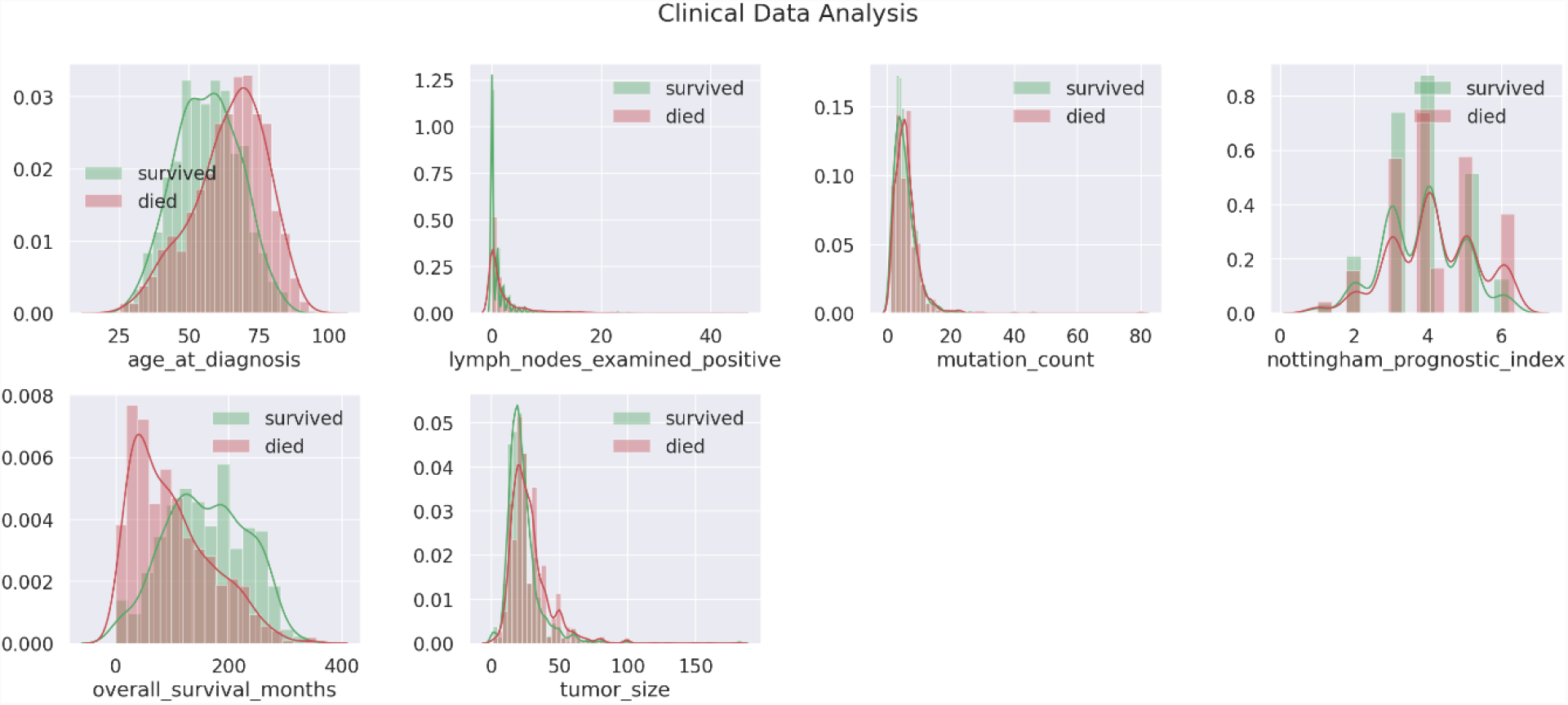
The Distribution in Numerical Clinical Columns

**Figure 3.**
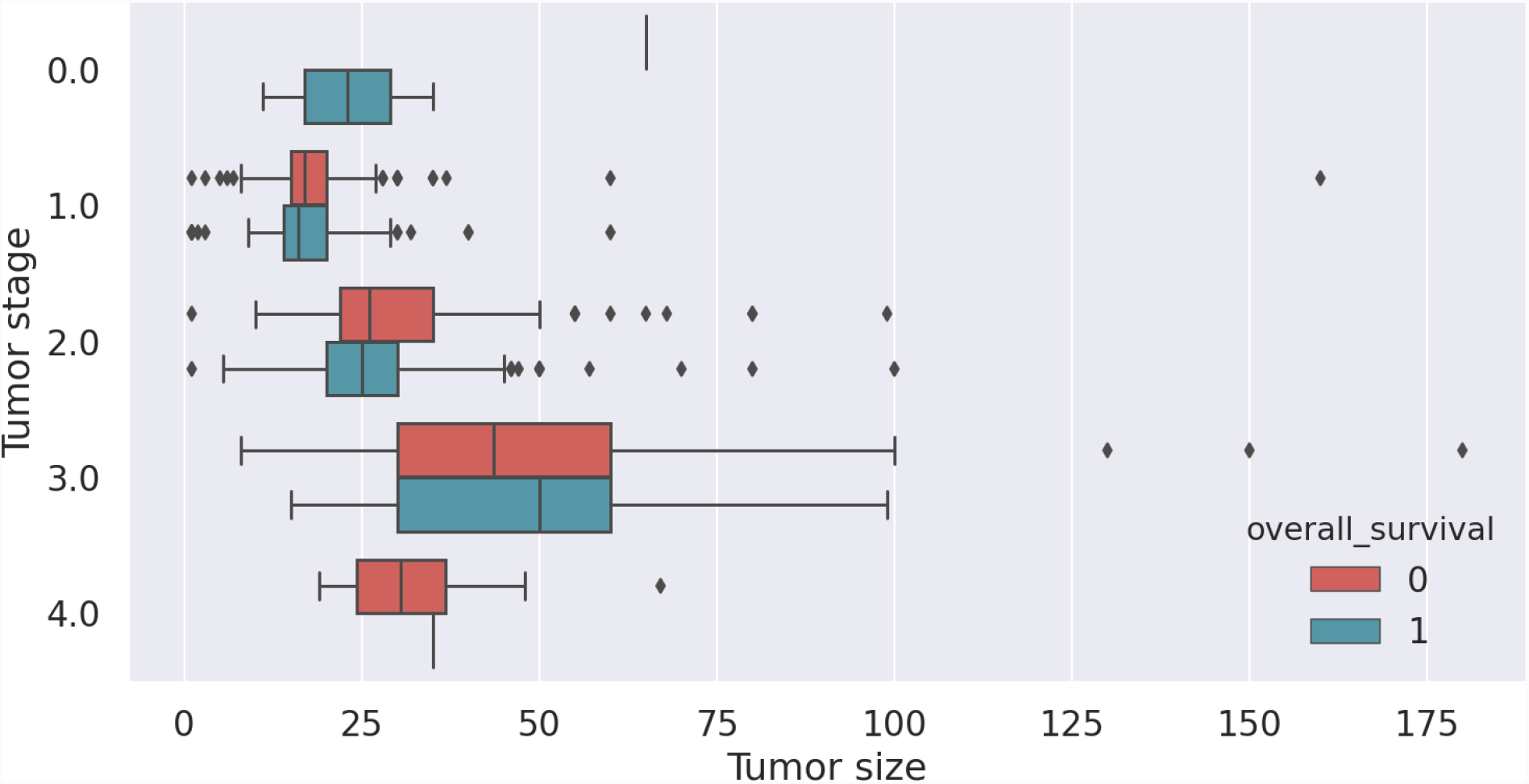
Tumor size

Another analysis that we can do in relation to probability distributions is to observe the age of the patients. you will notice that the average age of dead patients is closer to 75 years and the average age of people who survive the grip is 50 years

As the Tumer stage increases the tumor size increases as well. Also, if lower tumor stages the probability of survival is higher than when the patient reaches the fourth stage. you will notice that in green are the people who are alive and the size of the tumor, you will also notice in red the people who are dead in relation to the size of their tumor, the tumors are in a range from 0 to 50 in size.

The results (Figure 4) of the metrics related to modeling are quite significant. It is possible to observe mainly the accuracy of 79% of the levels of accuracy. You will also notice that if we only use genetic traits the results are not so good, which is why a greater variety of the dataset is important. CV scores: [0.75294118 0.78039216 0.74117647 0.76470588 0.78039216], CV Standard Deviation: 0.015369347405698364, CV Mean score: 0.76392156862745, Train score: 1.0, Test score: 0.794912559618442. The amount of true positives was between 316 and that was very good about Confusion Matrix.

[[316 55]

[74 184]]

**Figure 4.**
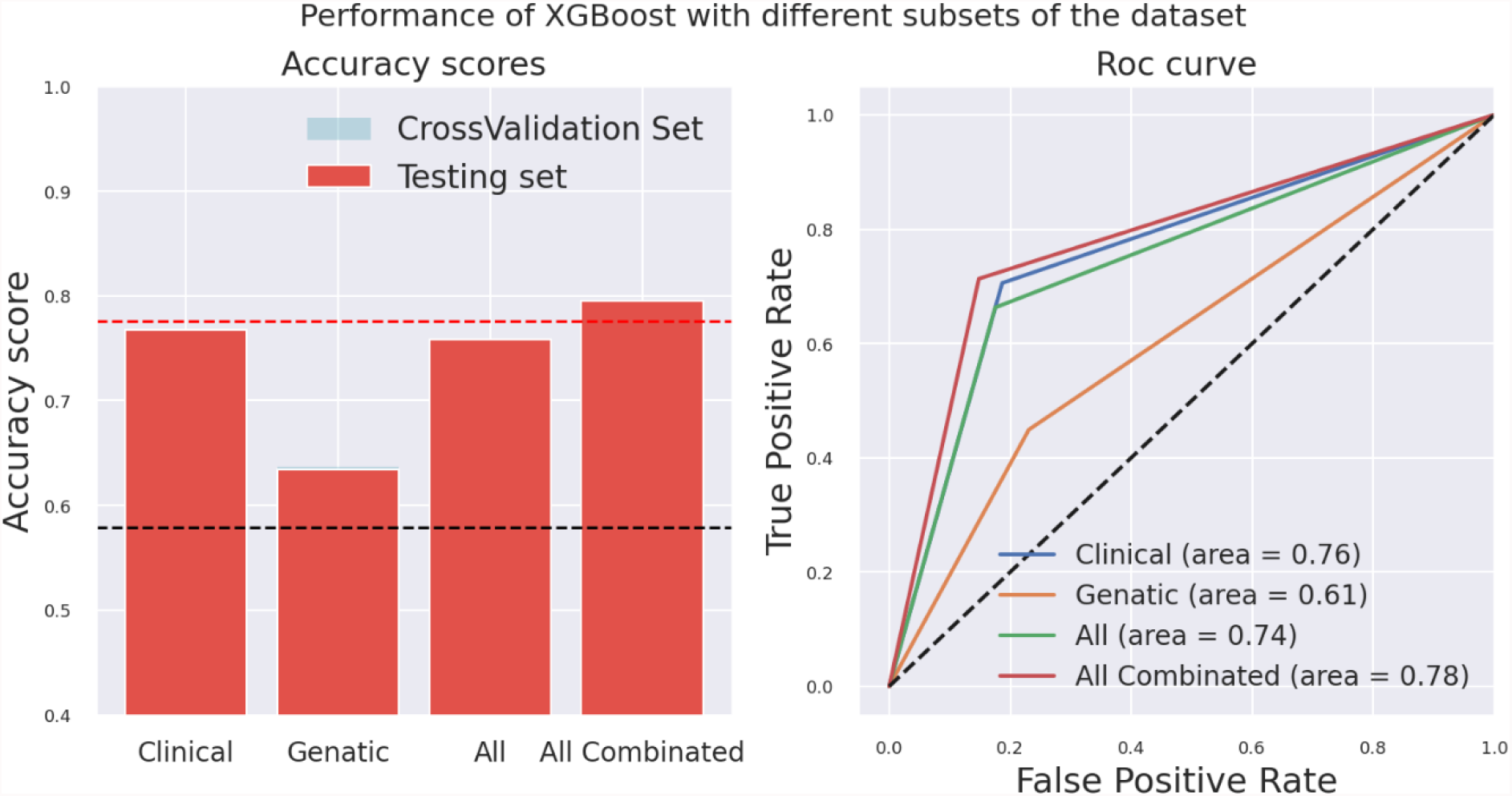
XGBoost preformed very well combared to traditional basic models

XGBoost preformed very well combared to traditional basic models, and the best model was the one that was trained with all of the features combined with accuracy of 0.779 and AUC of 0.76

## 5. Conclusion

The use of AI models on genetic data has great potential to improve our understanding of cancers and the prediction of survival. BigData open source datasets are available for researchers to analyze and hopefully get some interesting results. The best performing model was XGBoost with max_depth=5 and min_child_weight=1 which was trained with the complete dataframe with the addition of the entire combination of all genetic data values. The accuracy was 0.779 and the AUC was 0.76. To improve this project, scale up the data, include genetic mutations and raw genetic data in the modeling part, and do some deep learning models.

## Data Availability

All data produced are available online at cBioPortal

